# Genetic-Ancestry Analysis on >93,000 Individuals Undergoing Expanded Carrier Screening Reveals Limitations of Ethnicity-Based Medical Guidelines

**DOI:** 10.1101/2019.12.21.19015578

**Authors:** Kristjan E. Kaseniit, Imran S. Haque, James D. Goldberg, Lee P. Shulman, Dale Muzzey

## Abstract

**Purpose:** Despite strong association between genetic ancestry and carrier status, current carrier-screening guidelines recommend testing for a limited set of conditions based on a patient’s self-reported ethnicity, which conflates genetic and cultural factors.

**Materials and Methods:** For 93,419 individuals undergoing a 96-gene expanded carrier screen (ECS), correspondence was assessed among carrier status, self-reported ethnicity, and a dual-component genetic ancestry (e.g., 75% African/25% European) calculated from sequencing data.

**Results:** Self-reported ethnicity was an imperfect indicator of genetic ancestry, with 9% of individuals having >50% genetic ancestry from a lineage inconsistent with self-reported ethnicity. Self-reported ethnicity-based carrier-screening guidelines are incomplete, as several conditions not included in guidelines had similarly strong correlation between carrier rate and genetic ancestry as conditions included in screening guidelines. Limitations of self-reported ethnicity led to missed carriers in at-risk populations: for 10 ECS conditions, patients with intermediate genetic ancestry backgrounds—who did not self-report the associated ethnicity—had significantly elevated carrier risk. Finally, for seven of the 16 conditions included in current screening guidelines, most carriers were not from the population the guideline aimed to serve.

**Conclusion:** To provide equitable reproductive care, guidelines should discontinue the use of ethnicity as a basis for determining which patients are appropriate for carrier screening and instead recommend pan-ethnic ECS.

## Introduction

Carrier screening identifies couples at increased risk for conceiving fetuses affected by serious conditions that appreciably reduce lifespan, result in intellectual disability, and/or benefit from prenatal or perinatal intervention. Carrier screening is typically performed by first identifying female partners who are carriers for autosomal recessive and/or X-linked conditions, and subsequently testing their reproductive partners for condition(s) for which the female was a carrier.^1^ A couple is considered “at-risk” if the female carries a pathogenic variant that would cause an X-linked disease in a male child or if both partners are carriers of pathogenic variants in the same gene associated with an autosomal recessive disease.

Current professional guidelines by the American College of Medical Genetics and Genomics (ACMG) and the American College of Obstetricians and Gynecologists (ACOG) recommend pan-ethnic carrier screening for cystic fibrosis and spinal muscular atrophy.^2-4^ In addition, these two professional societies, as well as other organizations like the Jewish Genetic Disease Consortium (JGDC), have long recommended carrier screening for a partially overlapping and expanded set of conditions based on a patient’s self-reported ethnicity.

In the past decade, the evolution of carrier screening methodology (e.g., next-generation sequencing, “NGS”) has enabled scalable simultaneous screening of tens to hundreds of serious Mendelian diseases, termed expanded carrier screening (ECS). Pan-ethnic ECS has established analytical validity,^5^ clinical validity,^6^ clinical utility,^7^ and cost-effectiveness.^8^ Though ACOG considers ECS an “acceptable strategy” and both ACOG and ACMG generally acknowledge the complexities of race, ethnicity, ancestry, and coupling patterns in their carrier screening recommendations,^4,9-11^ both groups currently stop short of recommending that ECS be preferentially offered and instead continue to endorse self-reported ethnicity-based screening.

A patient’s self-reported ethnicity conflates genetic factors with cultural factors like traditions, lifestyle, diet, religious beliefs, and values,^12^ making self-reported ethnicity an indirect proxy for elevated carrier risk. A more direct relationship exists between carrier risk and genetic ancestry because both are based exclusively upon genetic variation. The analysis of genetic ancestry has been enabled by the genotyping of populations across the world (e.g., the Yoruba and Mandinka populations)^13,14^ from which allele frequencies of inferred ancestral populations (e.g., sub-Saharan African) are derived. An individual patient’s genetic ancestry can therefore be calculated by applying methods^15,16^ that ascertain the quantitative genetic ancestry proportion from each inferred ancestral population. In contrast, self-reported ethnicity is often qualitative, subjective, and ambiguous. For instance, 39.6% of individuals cannot identify the ancestry of all four grandparents,^17^ limiting the ability of self-reported ethnicity to reflect genetic ancestry. Further, the design of test requisition forms (e.g., binary choice vs. multiple categories) on which self-reported ethnicity is collected, and the stage at which the requisition is completed (i.e., pre-vs post-test), have been shown to influence self-reported ethnicity choice.^18,19^ The inability of self-reported ethnicity to accurately reflect genetic ancestry implies that ethnicity-based carrier screening recommendations may not achieve their intended effect, i.e., to identify those at highest risk of carrying serious conditions.

Genetic ancestry in the context of ECS was explored in a 2017 study of approximately 9,000 individuals.^19^ Here we employ a 10x-larger cohort to specifically evaluate the validity and impact of ethnicity-based carrier screening clinical guidelines. We analyzed the genetic ancestry of 93,419 individuals undergoing routine ECS for over 90 serious Mendelian disorders via NGS. We investigated the relationship between self-reported ethnicity and genetic ancestry, as well as the relationship between genetic ancestry and carrier status for the studied genetic disorders.

## Methods

### Study cohort

The study included individuals undergoing ECS (Foresight, Myriad Women’s Health) via NGS between 2014-01-01 and 2016-09-08 who had consented to anonymized research. The protocol for this study was reviewed and designated as exempt by Western Institutional Review Board and complied with the Health Insurance Portability and Accountability Act (HIPAA). Patients could select among the following self-reported ethnicity options on the test requisition form: Northern European, Southern European, French Canadian or Cajun, Ashkenazi Jewish, Mixed or Other Caucasian, East Asian, South Asian, Southeast Asian, African or African American, Hispanic, Middle Eastern, Native American, Pacific Islander, or Unknown. Patients choosing multiple self-reported ethnicities were counted as “Mixed or Other Caucasian,” and their multiple individual self-reported ethnicity selections (e.g., “Hispanic” and “Northern European”) were not accessible during the analysis. Patients with self-reported ethnicity of “Unknown” were excluded, as were those with self-reported ethnicity of “Finnish,” “Pacific Islander,” “Native American,” or “French Canadian/Cajun” due to insufficient sample size (<500 individuals in each subpopulation). An additional 621 individuals were excluded due to complex genetic ancestry (see below), resulting in a final analyzed cohort of 93,419 individuals.

### Carrier screening

All patients were analyzed with short-read NGS of exons and certain intronic regions, as well as a specialized assay for fragile X syndrome.^5^ DNA was extracted from blood or saliva. The 96 diseases and corresponding genes included for analysis are listed in **SI Table 1**. Conditions for which there are multiple manifestations of disease severity (e.g., classical (severe) and non-classical (moderate) forms of 21-hydroxylase-deficient congenital adrenal hyperplasia) were analyzed as a single, combined condition. A carrier was defined as a patient harboring a pathogenic or likely pathogenic variant in a gene of interest. Certain high-frequency variants or variant combinations involving complex interpretation or mild phenotypes—e.g., the NM_000500.7(CYP21A2):c.955C>T(Q319*) variant in combination with a duplication for congenital adrenal hyperplasia (CYP21A2-related), or the NM_000060.2(BTD):c.1330G>C(D444H) variant for biotinidase deficiency—were considered benign for population-level carrier rate analyses and are enumerated in **SI Table 2**.

When analyzing guidelines, we took an inclusive approach: if either ACMG or ACOG recommended that patients should be screened or that screening should be considered for a disease, we considered the disease to be part of current guidelines (**SI Table 3**).

### Genetic ancestry analysis

On- and off-target reads obtained during ECS were used for genetic ancestry analysis after read mapping quality and base-quality-score filters were applied, as described by Wang *et al*.^20^Genetic ancestry was inferred using a proprietary analysis method (patent application WO2018144135A1). In brief, the method is based on sparse non-negative matrix factorization (with seven components) and was used to estimate ancestry shared with hypothetical ancestral populations using an imputed reference panel of 51 populations^13^ and an additional imputed reference panel of the Ashkenazi Jewish population (GEO accession no. GSE23636).^21^ Ancestry was estimated using distances along pair-wise ancestry clines between reference populations (e.g., a European-African cline): each patient was assigned a score from 0 (0%) to 1 (100%) for each genetic ancestry component, with at most two non-zero components. The genetic ancestry components determined were (sub-Saharan) African, Ashkenazi Jewish, European, Middle Eastern, South Asian, East Asian, Native American, and Pacific Islander. For finer resolution of genetic ancestry among patients with >87.5% South Asian, Middle Eastern, Ashkenazi Jewish or European genetic ancestry from the analysis using the first reference panel, we used a second reference panel containing only those populations (four components). 621 individuals (0.66%) could not be confidently categorized as belonging to a single genetic ancestry cline between two populations, mostly due to ancestry from three or more sources (e.g., African, European, and Native American genetic ancestry). Individuals who were not included in subsequent analyses mostly self-reported as Hispanic (506, 81%), “Mixed or Other Caucasian” (71, 11%), and African or African-American (32, 5%), with the remaining 12 individuals characterized by other self-reported ethnicities.

### Matching populations among guidelines, self-reported ethnicity, and genetic ancestry

The ethnicity categories used in guidelines and in the ECS test requisition forms do not match perfectly (e.g., a patient can self-report as Southern European, but this is not an ethnicity category for ACOG or ACMG). Furthermore, due to constraints of the ancestry analysis, not all self-reported ethnicities for which there are guidelines were associated with a single genetic ancestry component. The correspondences among self-reported ethnicities and genetic ancestries included in ACOG/ACMG guidelines, the test-requisition forms, and our genetic ancestry analysis are listed in **SI Table 3**.

### Categorizing genetic ancestry as low, medium, or high

For all ancestries, we defined a “low” amount of genetic ancestry as being less than 1/32. We then considered genetic ancestry in increments of 1/32 and defined a threshold for a “high” amount of genetic ancestry (*g*) based on the smallest range of genetic ancestry—*g*…*g* + 1 / 32—in which more than half of individuals self-reported the associated ethnicity. For instance, African genetic ancestry between 15/32 and 16/32 was the lowest genetic ancestry range in which over 50% of patients had a self-reported ethnicity of African or African-American; thus, *g*=15/32 was the threshold between “high” and “medium” African genetic ancestry. We defined a “medium” amount of genetic ancestry as being between the two threshold values (i.e., more than “low” threshold of 1/32 but less than the “high” threshold at *g*). The threshold for European genetic ancestry was determined based on both Northern and Southern Europeans.

## Results

### Self-reported ethnicity is an imperfect indicator of genetic ancestry

We quantified the agreement between self-reported ethnicity and genetic ancestry by categorizing individuals according to the source of the genetic ancestry component responsible for >50% of their ancestry (“GA_majority_”). Overall, 9% of patients had a GA_majority_ that was unexpected based on the self-reported ethnicity (excluding those self-reporting as “Mixed or Other Caucasian”). Concordance between self-reported ethnicity and GA_majority_ was highest in those who self-reported as Northern European (96.9%), Hispanic (96.4%), South Asian (96.3%), or Southeast Asian (95.8%) (**Figure 1a**). Concordance was lowest (<90%) in those self-reporting as Middle Eastern (59.2%), Ashkenazi Jewish (80.2%), or Southern European (84.0%) (**Figure 1a**). We also observed wide distributions of the genetic ancestry proportions within self-reported ethnicity groups both above and below the 50% threshold (**Figure 1b**), suggesting that genetic ancestry level can vary among individuals in the same self-reported ethnicity and that substantial genetic ancestry may still be present even if GA_majority_ and self-reported ethnicity do not match. Nearly one-third of patients (31%) had an self-reported ethnicity of “Mixed or Other Caucasian” (either by directly selecting this option on the requisition form or by selecting more than one ethnicity; see Methods), with most having GA_majority_ of European (**Figure 1a**, “Other” row). **Supplemental Text 1** describes these and further observations in more detail.

**Figure 1.**
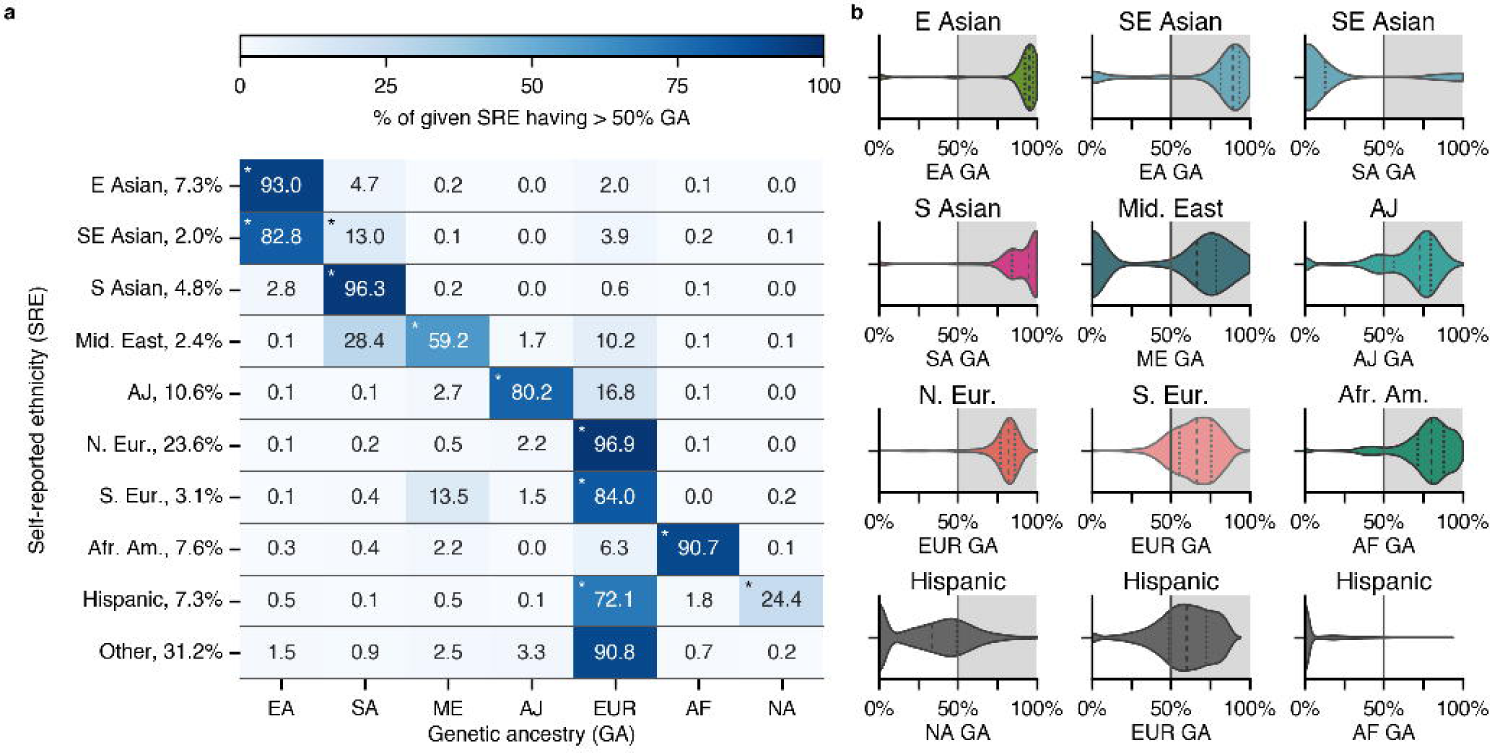
Imperfect agreement between self-reported ethnicity and genetic ancestry. **(a)** Individuals were categorized according to the source of the genetic ancestry component responsible for >50% of their ancestry (GA_majority_). Each row corresponds to a self-reported ethnicity group and sums to 100% (Pacific Islander genetic ancestry not displayed). “Expected” genetic ancestry roughly corresponds to the diagonal and is marked with asterisks (*). **(b)** Distribution of genetic ancestry (subpanel x-axis) among self-reported ethnicity groups (subpanel titles). Vertical lines within violin plots indicate the 25th, 50th, and 75th percentiles of genetic ancestry. The highlighted region to the right of the vertical 50% line indicates agreement between self-reported ethnicity and GA_majority_. *EA*: East Asian, *SA*: South Asian, *ME*: Middle Eastern, *AJ*: Ashkenazi Jewish, *EUR*: European, *AF*: African, *NA*: Native American genetic ancestry.

### Assessing carrier rates as a function of genetic ancestry

Because it has multiple high-frequency recessive conditions and historically high uptake of genetic screening, the Ashkenazi Jewish population has been extensively addressed by ACMG, ACOG, and JGDC carrier screening guidelines. Of the three, JGDC has the largest recommended panel. To assess the validity and completeness of these panels, we analyzed the relationship between carrier rates and the amount of AJ genetic ancestry for diseases on the JGDC Ashkenazi panel, as well as for diseases outside this panel. As expected, we observed that diseases on the ACOG, ACMG, and JGDC Ashkenazi panels were largely enriched in the AJ self-reported ethnicity group and displayed an increasing carrier-risk profile as a function of AJ genetic ancestry (**Figure 2a & 2e**). Diseases not on the JGDC panel mostly displayed “flat” carrier-risk profiles (i.e., AJ genetic ancestry had no strong effect on the carrier rate) and no strong enrichment in the AJ self-reported ethnicity group (**Figure 2b**). Diseases that have a strong genetic ancestry association with an ancestry other than AJ showed a downward-sloping carrier risk-profile as a function of AJ genetic ancestry: more AJ genetic ancestry meant less of the risk-associated genetic ancestry, thereby reducing the carrier risk (**Figure 2c**). Notably, we also observed several diseases not on the JGDC panel that were enriched in the AJ self-reported ethnicity group and showed an increasing carrier-risk profile as a function of AJ genetic ancestry (**Figure 2d & 2e**), suggesting that panels intended to screen for conditions enriched in the AJ population are incomplete.

**Figure 2.**
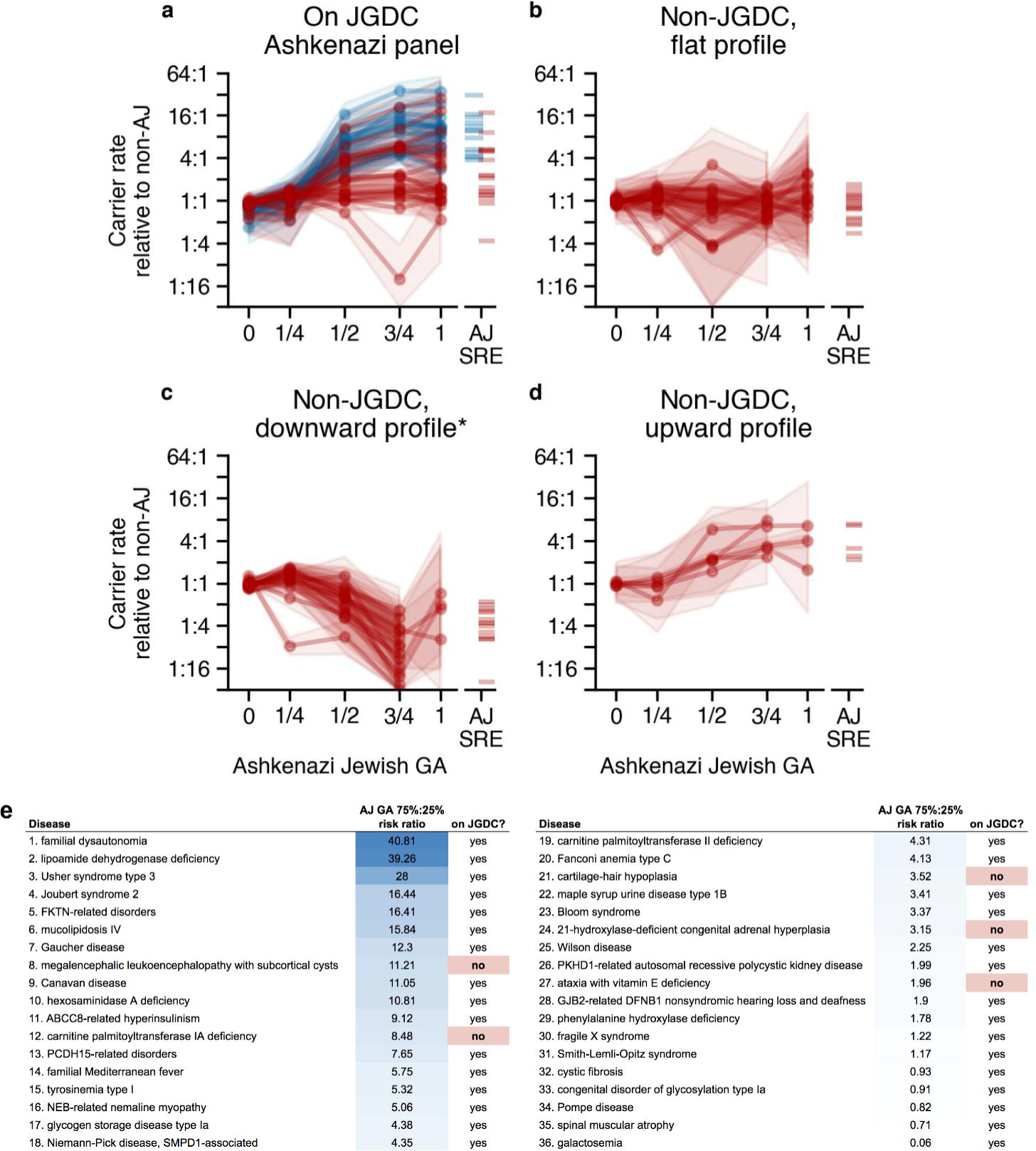
Ashkenazi Jewish (AJ) guideline recommendations lack several diseases for which AJ patients have elevated carrier risk. Each trace corresponds to an ECS panel condition, and the y-axis shows the ratio of carrier rates for the group of patients with a genetic ancestry indicated on the x-axis (numerator) relative to the group of patients who did not self-report as AJ (denominator). The right of each panel computes the same ratio where the numerator is the carrier rate among patients who self-reported as AJ. Diseases highlighted in blue in **(a)** are also part of screening guidelines from ACMG and/or ACOG. To classify diseases as “flat” **(b)**, “downward” **(c)**, or “upward” **(d)**, the ratio of y-axis values at genetic ancestries (x-axis values) of 3/4 (numerator) and 0 (denominator) was calculated. A “downward” profile had ratio < 0.5; a “flat” profile had 0.5 ≤ ratio < 2, and an “upward” profile had both a ratio ≥ 2 and a carrier rate relative to non-AJ > 1 for genetic ancestry of 3/4. Diseases not meeting these criteria were not categorized. Error boundaries represent the 95% confidence interval calculated from a z-test on the log-transformed ratios. **(e)** Listing of conditions on the JGDC panel or that show enrichment in the AJ population based on the carrier-rate ratio of at 75% and 25% from panel **(d)**. * only 15 diseases shown for **(c)**.

### Carrier risk enrichment in those with substantial, but unreported ancestry

After observing gradation in carrier risk as a function of genetic ancestry in the AJ population that cannot be fully captured by self-reported ethnicity, we explored whether the limitations of self-reported ethnicity would also be expected to affect carrier identification in other populations. Specifically, we hypothesized that individuals with medium genetic ancestry who did not self-report the corresponding ethnicity may have an increased carrier risk, yet their elevated risk would not be captured by ethnicity-specific guidelines. For each self-reported ethnicity (SRE), we first partitioned individuals into +SRE (i.e., selected the ethnicity on the requisition form) and -SRE (i.e., did not select the ethnicity) groups, and then further subdivided the groups based on the corresponding genetic ancestry, resulting in high, medium, and low genetic ancestry subgroups (**Figure 3**; see Methods).

**Figure 3.**
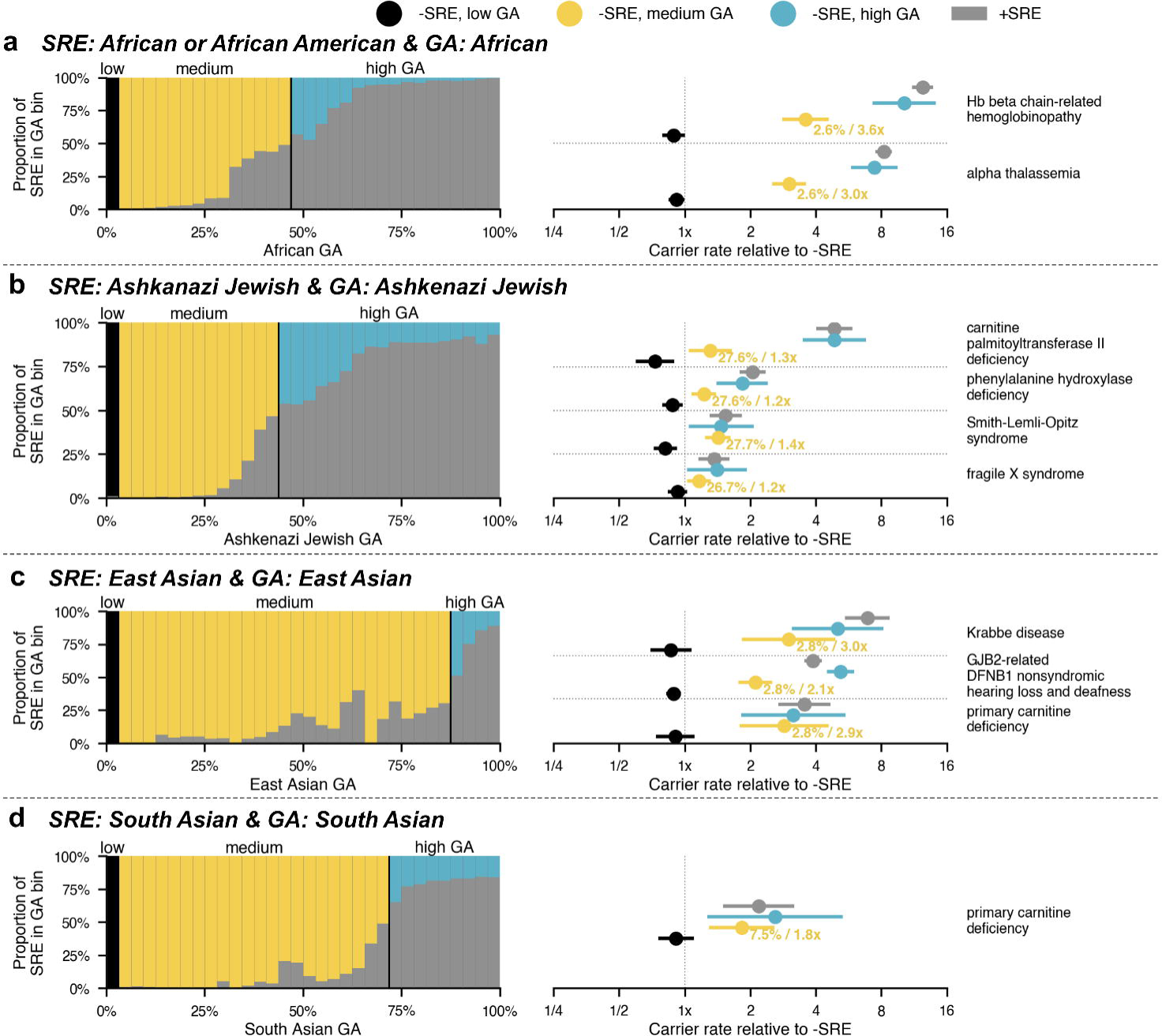
For several conditions, patients who have medium genetic ancestry and do not self-report the associated ethnicity have significantly elevated carrier risk. For progressively higher levels of genetic ancestry (in 32 bins from 0% to 100% on the x-axis), the left panels of **(a)**-**(d)** indicate the relative proportion of patients who self-report the associated ethnicity (see key at top for color scheme). The cutoff between medium genetic ancestry and high genetic ancestry is described in Methods. For the indicated conditions, panels on the right show the relative carrier rate of patients in each sub-cohort (colored as in the left panels). Percentages shown in yellow in the right panel indicate the proportion of non-self-reporting individuals with medium genetic ancestry screened for the given disease, with an additional annotation indicating the relative carrier risk value. A condition is plotted if the carrier rate was significantly greater in (1) the +SRE population, (2) the -SRE medium-genetic ancestry population, and (3) the -SRE high-genetic ancestry population, compared to the -SRE population based on the 95% confidence interval of the risk ratio (estimated using a Z-test of the log-transformed risk ratio). While cystic fibrosis met these criteria in the Northern European population, it is recommended for pan-ethnic screening by both ACMG and ACOG and was thus omitted from the figure.

We observed that a substantial proportion of individuals fell into the “medium genetic ancestry” subgroup. For example, while 7,100 (7.6%) patients in the entire cohort self-reported as African or African-American, 2,800 (3.0%) other patients had medium African genetic ancestry and did not self-report as being African or African-American. The latter group would not have been screened under ethnicity-specific guidelines, yet they had a 3.6-fold higher risk of being a carrier of Hb beta chain-related hemoglobinopathy and a 3.0-fold higher risk for being a carrier of alpha thalassemia relative to -SRE individuals (**Figure 3a**). Among all patients with medium African genetic ancestry, just 16% self-reported as African or African-American; the others tended to self-report as Hispanic (47%) or “Mixed or Other Caucasian” (25%) (**SI Figure 2**), meaning that self-reported ethnicity-based guidelines would not have captured their elevated risk for being carriers of hemoglobinopathies.

Importantly, the insufficiency of self-reported ethnicity as a means to identify elevated carrier risk for particular conditions was true for other ancestries as well, including the AJ (**Figure 3b**), East Asian (**Figure 3c**), and South Asian (**Figure 3d**) populations. Our observation of carrier-rate enrichment among -SRE patients with medium genetic ancestry in **Figure 3** would be trivial if it were composed of “Mixed or Other Caucasian” patients where one of their several reported ethnicities was the ethnicity of interest (See Methods); however, we observed comparable carrier-rate enrichment among -SRE patients with medium genetic ancestry even when we altogether excluded “Mixed or Other Caucasian” patients (**SI Figure 2**). These data suggest that many patients had elevated carrier risk due to their genetic ancestry and did not self-report an ethnicity that would have indicated their risk.

Patients with high levels of genetic ancestry did not always self-report the associated ethnicity, and carrier rates were comparable irrespective of self-reporting (blue vs gray, **Figure 3**). When patients did not self-report the ethnicity associated with their high genetic ancestry, they typically self-reported as “Mixed or Other Caucasian” or as a related population (e.g., those with high East Asian genetic ancestry self-reporting as Southeast Asian) (**SI Figure 3**). Although we do not know why the patients in our cohort reported an ethnicity discordant with their high genetic ancestry, they nonetheless may not have received screening appropriate for their risk if ethnicity-based guidelines had been strictly followed.

### Both ethnicity- and ancestry-based guidelines miss substantial proportions of carriers

We hypothesized that carriers of conditions with ethnicity-specific guidelines could be missed because of deficiencies in self-reporting (i.e., those with medium or high genetic ancestry do not self-report the related ethnicity) and/or because many carriers exist outside of the presumed high-risk ancestries (i.e., an abundance of carriers in the low-genetic ancestry group, which has many patients and a low carrier rate).

We observed that within the +SRE group, most carriers for conditions in current ACOG and ACMG guidelines had medium or high genetic ancestry, while in the -SRE group the proportion of carriers from the low genetic ancestry group was substantial for all diseases (**Figure 4**). This suggests that a large proportion of carriers would still be missed if current ethnicity-based guidelines were adapted to be ancestry-based instead (**Figure 4**, comparing the hatched black regions to all other regions). By contrast, pan-ethnic carrier screening would identify all carriers, regardless of their level of genetic ancestry or self-reported ethnicity.

**Figure 4.**
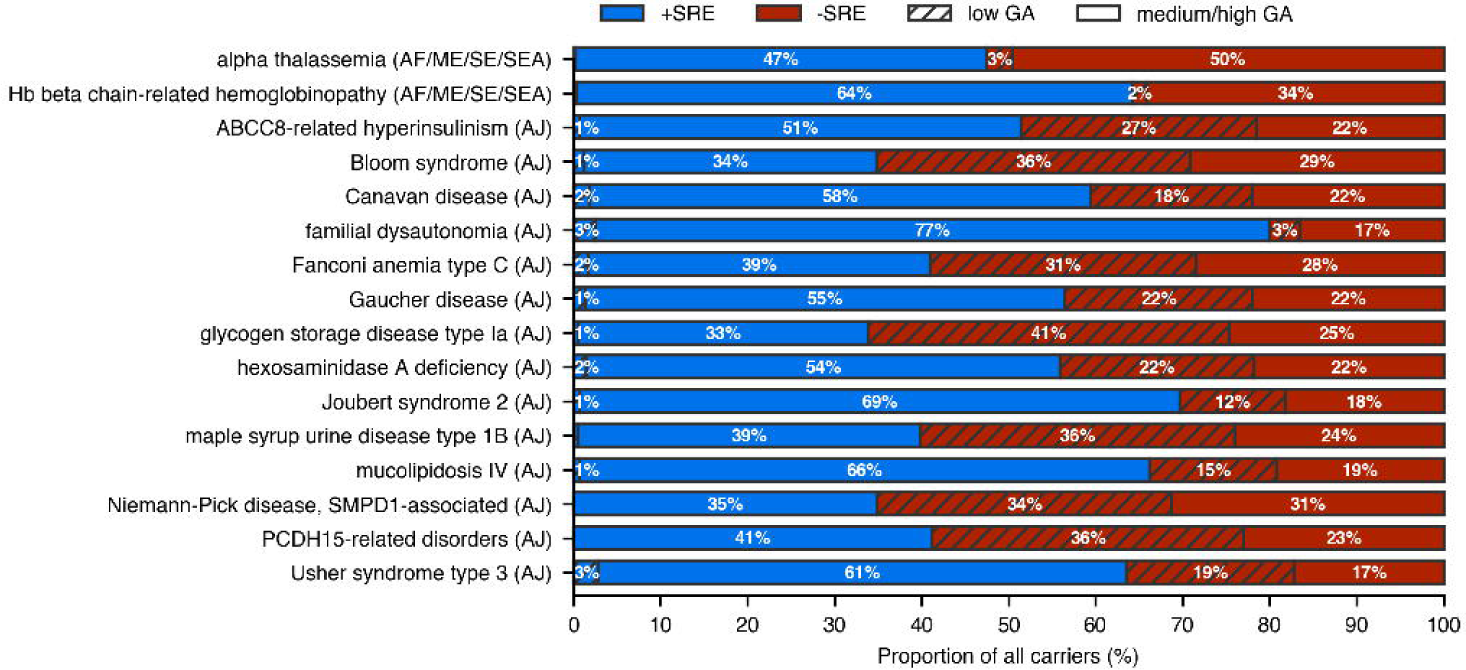
Distributions of carriers for diseases in the ACMG or the ACOG guidelines in the +SRE vs. -SRE (blue vs red) and low vs medium/high genetic ancestry (hatched vs unhatched) groups show that both ethnicity- and ancestry-specific screening miss a substantial proportion of carriers. For diseases with multiple guideline-ethnicities (alpha thalassemia and Hb beta chain-related hemoglobinopathies), a person was considered in the “low genetic ancestry” group if all the related ancestries (shown in parentheses on each row) were low (<1/32). For the Southeast Asian guidelines, determination of low vs high/medium genetic ancestry was based on South Asian genetic ancestry instead of East Asian genetic ancestry (giving a smaller proportion of carriers in the -SRE, low genetic ancestry group). For seven out of the 16 diseases, most carriers were in the -SRE population; for seven diseases, at least 25% of carriers were in the low genetic ancestry population. Cystic fibrosis and spinal muscular atrophy were not included, as pan-ethnic carrier screening is recommended for these diseases. *EA*: East Asian, *SA*: South Asian, *ME*: Middle Eastern, *AJ*: Ashkenazi Jewish, *EUR*: European, *AF*: African, *NA*: Native American genetic ancestry.

The distributions of frequencies for variants observed in the low vs. medium/high genetic ancestry groups differed, with each group typically having some distinct variants and the medium/high genetic ancestry group having relatively less variant diversity (as expected in the case of founder or ancestry-related variants) (**SI Figure 4**). These data suggest that targeted genotyping carrier screening that interrogates only a subset of variants (as opposed to novel variant detection via NGS) would miss carriers outside of the target population.

Overall, ethnicity-based guidelines would have missed 77% of carriers in the study cohort, with large differences per ethnicity (**Figure 5, SI Table 4**). Nearly one-third (32%) of carriers for Mendelian diseases among African-Americans would have been missed based on screening guidelines from ACMG and ACOG; that number rises to 87% of carriers missed among Hispanics when guidelines are followed. Missed carriers are associated with 69 to 94 distinct diseases, depending on the ethnicity (**SI Table 4**). Furthermore, depending on the ethnicity, only 7%-49% of guideline-missed carriers were accounted for by a single disease (**SI Table 4**), suggesting that the incremental addition of single diseases to current guidelines is not sufficient to close the gap in carrier identification. An ancestry-specific screening approach would have a marginal improvement over ethnicity-specific carrier screening in some ethnicities, yet it would perform worse in other ethnicities (red vs yellow bars, **Figure 5**). A pan-ethnic carrier screening approach would identify the carriers of diseases currently in guidelines but would miss carriers for all other diseases (gray vs teal bars, **Figure 5**). These data suggest that current guidelines do not identify most carriers for serious recessive diseases, and that either pan-ethnic ECS (teal bars, **Figure 5**) or the addition of many disease-ethnicity pairs to current guidelines would be needed to adequately identify carriers in the population.

**Figure 5.**
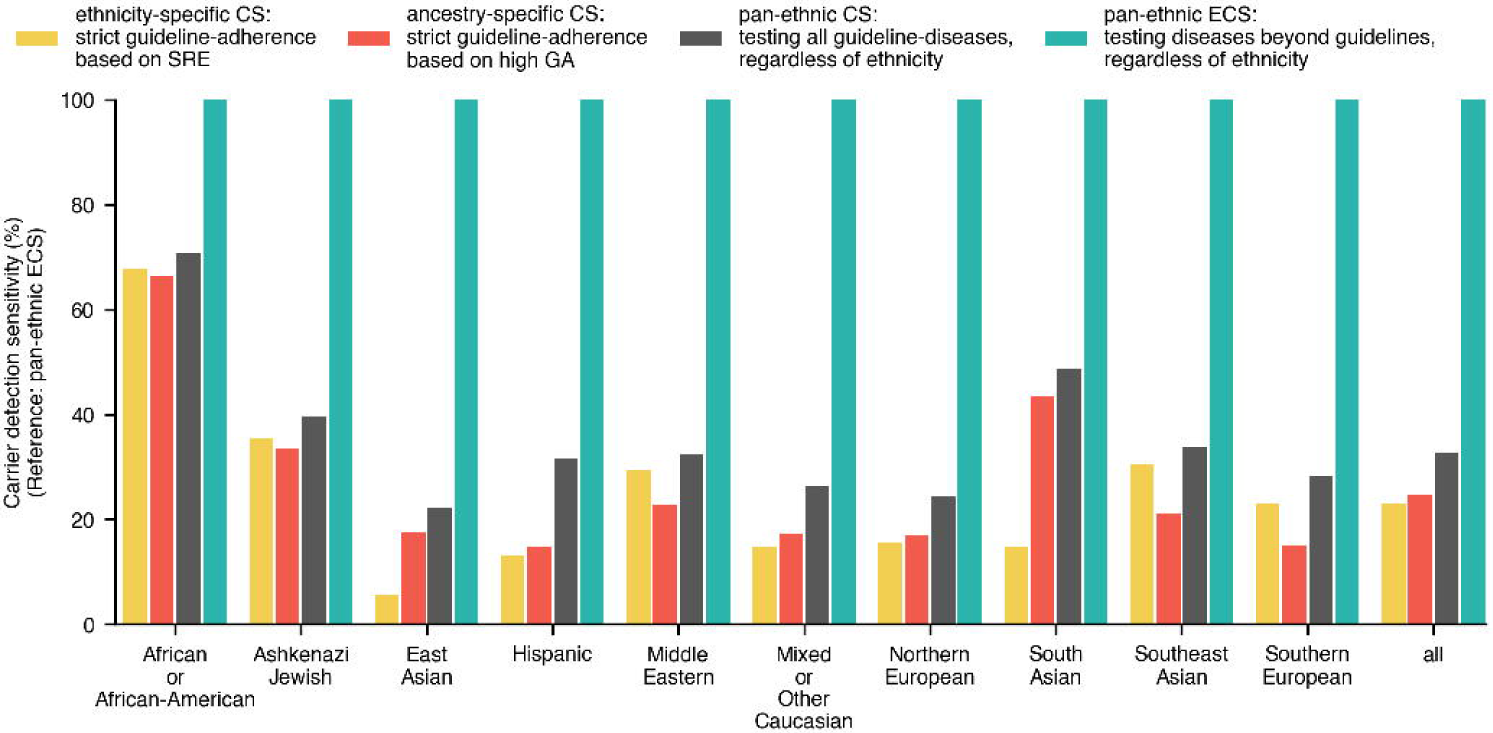
Comparison of carrier-detection sensitivity for different carrier screening scenarios across SREs. Ethnicity-specific and ancestry-specific carrier screening include pan-ethnic screening of cystic fibrosis and spinal muscular atrophy. Ancestry-specific carrier screening considers a person eligible for testing if they have “high” levels of any relevant genetic ancestry for a given guideline-disease.

## Discussion

Here we evaluated the validity of current carrier screening guidelines by exploring the relationship between self-reported ethnicity, genetic ancestry, and carrier status for a large cohort tested with a 96-gene ECS panel. For the following reasons, we conclude that current ethnicity-based guidelines have consequential deficiencies that impact patient care. First, our data demonstrate that self-reported ethnicity is an imperfect indicator of genetic ancestry, which is problematic if the former guides who is offered screening but the latter guides who is a carrier. The gradation of risk by genetic ancestry and the wide distribution of genetic ancestry within a self-reported ethnicity group suggests that the binary nature of self-reported ethnicity may be too coarse to capture clinically meaningful risk enrichment. Second, our analysis revealed that ethnicity-specific guidelines are incomplete, as multiple diseases were enriched in a well-studied population (Ashkenazi Jewish) but missing from that population’s guidelines. Third, even if ethnicity-specific guidelines were revised to recommend more diseases in particular ethnicities, carriers would still be missed because many individuals with elevated levels of a genetic ancestry do not self-report the associated ethnicity. Fourth, for seven of the 16 diseases in guidelines, the majority of the carriers identified in our cohort were outside the population with a guideline. In several cases, the risk was not due to self-reporting limitations but rather to the non-zero carrier rate in non-guideline populations. Pan-ethnic screening—currently deemed acceptable but not yet recommended by medical societies—overcomes these limitations of ethnicity-based screening.

In past decades, when carrier screening panels were costly to offer and challenging to expand with content, ethnicity-specific screening could provide efficient allocation of health-care resources by screening for a limited set of diseases in high-risk populations. This approach had important successes: ethnicity-based screening for Tay-Sachs disease in the AJ population has reduced the number of affected pregnancies in that population so precipitously that there are now relatively more Tay-Sachs disease-affected pregnancies in non-AJ ethnicities.^22^ But, for several reasons, the original motivations for limiting the recipients and content of carrier screening panels are no longer compelling. First, risk is typically not concentrated in specific ethnicities and is being further spread among populations due to changing demographics and increased inter-group marriage. This increased admixture is reflected in our results: 31% were “Mixed or Other Caucasian,” and genetic ancestry varied widely in certain self-reported ethnicity groups. Second, it is not clear that ethnicity-based guidelines are efficient, as a previous study found that an average of 2-5 minutes of office visits were consumed with selection of the ethnic origins of patients undergoing prenatal sickle cell screening.^18^ Third, Condit et al.^17^ showed that a large proportion of patients cannot accurately identify the ethnicities of their ancestors. And fourth, the cost of screening for additional conditions is now marginal due to advancements in DNA sequencing methodologies. These factors suggest that modern pan-ethnic ECS panels are an efficient means to assess reproductive risk for recessive disease.

Our data show that challenges would remain even if a genetic ancestry-based approach, rather than the current self-reported ethnicity-based approach, were used to administer carrier screening. Such genetic ancestry-based guidelines (e.g., screen for condition X if the patient has ancestry for population Y above threshold Z) may still perpetuate the inequities and stigmatization associated with ethnicity-based screening because they restrict screening to only certain populations.^23,24^ It is also not clear how a genetic ancestry-based carrier screening approach could be efficiently executed on a population scale. For example, if genetic ancestry assessment preceded carrier screening, detailed specifications would be needed to describe how to transmit a patient’s genetic ancestry information to carrier screening laboratories and how the resulting panels should be selected. If genetic ancestry assessment were performed by the carrier screening laboratory, the panel-selection challenges would be compounded by ethical issues, such as potentially needing to notify patients of unexpected genetic ancestry in their lineage. For these reasons, as well as results from this study showing that a genetic ancestry approach would have suboptimal carrier detection in some ethnicities, we submit that pan-ethnic ECS is the most effective strategy to maximize carrier identification and simplify administration of population-wide carrier screening. However, using genetic ancestry as a means to calculate patient-tailored residual risk upon screening negative may have promising potential.

Our results generally provide support for the broad adoption of pan-ethnic ECS, but we have not directly explored herein the potentially negative consequences of pan-ethnic ECS, such as increased provider burden for partner testing, reduced perceived clinical utility for screening conditions not historically included in guidelines, and reduced cost effectiveness. However, these possible consequences have been explored and refuted elsewhere. First, addition of more carrier screening conditions to a panel does not linearly increase the frequency with which a carrier’s partner would need to be tested: on a 176-condition panel, pan-ethnic screening of just the 18 (10.2%) most prevalent conditions would account for 61% of carriers and 84% of at-risk couples detected by the entire panel.^25^ Second, in a clinical utility study of ECS, patients’ pursuit of alternative reproductive options was driven largely by the severity of the condition for which their children were at risk, not by whether the condition was part of guideline-based screening; indeed, 72% of the at-risk couples who planned or pursued an alternative reproductive option were carriers for conditions outside of guidelines.^7^ Finally, due to life-years gained from screening for a broader array of conditions, a 176-gene panel showed higher cost-effectiveness at multiple price points relative to screening for only cystic fibrosis (by genotyping) and spinal muscular atrophy.^8^

The results of this study could be further refined in the future. For instance, the set of conditions and populations analyzed could be expanded, empowering genetic ancestry analysis that interrogates sub-ancestries with greater precision. In particular, the delineation of individuals with joint European-South Asian ancestry versus those with joint European-Middle Eastern ancestry could be improved upon (miscategorizing these ancestry clines could partially underlie the 28% of self-reported Middle Easterners with South Asian GA_majority_; the reversed combination—i.e., high proportion of self-reported South Asians with Middle Eastern GA_majority_— was not observed). Further investigation is also warranted for individuals reporting multiple ethnicities on the requisition form, who were considered as belonging to the “Mixed or Other Caucasian” self-reported ethnicity group. While our findings indicate that over 90% of these patients have European GA_majority_, factors influencing the choice of multiple ethnicities are of interest but could not be investigated here.

Taken together, our observations—coupled with studies demonstrating the analytical validity, clinical validity, clinical utility, and cost effectiveness of ECS—elucidate the merit of recommending pan-ethnic ECS rather than perpetuating ethnicity-specific guidelines that yield inequities in reproductive care and limit the impact of carrier screening.

## Data Availability

Data will be shared as permitted upon reasonable request to the authors.

## Acknowledgements

The authors would like to thank the patients that made this study possible. We would also like to thank and acknowledge Katie Johansen Taber and Aishwarya Arjunan for their thorough comments on our manuscript.

## Figure Legends

**Supplemental Figure 1**. *Distribution of genetic ancestry in all self-reported ethnicity groups*. Highlighted violin plots indicate the “expected” genetic ancestry for the given self-reported ethnicity. Each violin plot includes only those with ≥1/32 of the given genetic ancestry, with the numbers indicating the proportion of the self-reported ethnicity plotted as well as the absolute number. Subpopulations with fewer than 100 individuals were not shown. Vertical lines within the violin plot indicate the 25th, 50th, and 75th percentiles of genetic ancestry. The “any” self-reported ethnicity includes the whole study cohort.

**Supplemental Figure 2**. *Same as Figure 3 but excluding “Mixed or Other Caucasian” patients*.

**Supplemental Figure 3. (a and c)** Distribution of self-reported ethnicities among those with medium genetic ancestry (a) and high genetic ancestry (c). Hatching indicates secondary self-reported ethnicities. Colors are indicated in panel (c), where the text overlay specifies the ethnicity indicated by the leftmost bar. **(b and d)** Comparison of ancestry-specific carrier rates among those with medium genetic ancestry (b) and high genetic ancestry (d) based on whether they self-reported the associated ethnicity (x-axis) or not (y-axis). Each dot represents a combination of a disease and an ancestry (e.g., cystic fibrosis in Europeans). Disease-ancestry-self-reported ethnicity combinations with no observed carriers in this dataset are shown on the axes.

**Supplemental Figure 4**. *Variant frequency distribution in those with low vs medium or high guideline-related genetic ancestry*. For the 16 conditions with ethnicity-specific guidelines (separate panels), patients with low genetic ancestry (left of panel) and medium or high genetic ancestry (right of panel) carry pathogenic variants at different frequencies (y-axis). The genetic ancestry was chosen to correspond to the ethnicity with a guideline (e.g., Ashkenzi Jewish for Gaucher disease). Each horizontal line represents a variant, with a blue line indicating a variant observed in only one of the two subpopulations (e.g., in the low genetic ancestry subpopulation but not in the medium or high genetic ancestry subpopulation) and a black trace spanning the subpopulations indicating a single variant present in both subpopulations often at different frequencies (hence the upward or downward slope of middle segment). For most conditions, the medium-or-high-genetic ancestry subpopulation exhibited smaller variant diversity (i.e., fewer horizontal lines on right side of each panel) compared to the low-genetic ancestry subpopulation, as expected if founder variants are present.

**Supplemental Table 1**. *Diseases included in the analysis*.

**Supplemental Table 2**. *Variants excluded in carrier rate analysis*.

**Supplemental Table 3**. *Self-reported ethnicities, genetic ancestries, and guideline recommendations as analyzed in this study*.

**Supplemental Table 4**. *Carriers missed by ethnicity-specific carrier screening compared to pan-ethnic expanded carrier screening*. Individuals carriers for multiple diseases were counted for each disease.

**Supplemental Text 1**. *Observations of unexpected majority-genetic ancestry*.

The test requisition form for our ECS lists three Asian ethnicity options (East, Southeast, and South Asian) and lists example countries of origin accompanying each option. We observed that those self-reporting as Southeast Asian typically had their majority genetic ancestry coming from the East Asian (EA) rather than South Asian (SA) component (**Figure 1a**, “E Asian”, “SE Asian” and “S Asian” rows). There were both East Asians with majority South Asian genetic ancestry (4.7% of self-reporting East Asians) as well as South Asians with majority East Asian genetic ancestry (2.8% of South Asians). The largest proportion of individuals with majority-European ancestry (EUR) among the three Asian SREs was among the Southeast Asian group (3.9% of self-reporting Southeast Asians, compared to 2.0% of East Asians and 0.6% of South Asians). The majority genetic ancestry and self-reported ethnicity matched better for South Asians (96.3% of South Asians had majority genetic ancestry component of South Asian ancestry) compared to East Asians (93.0%).

Those self-reporting as Middle Eastern had majority genetic ancestry split mostly between Middle Eastern (59.2%, ME), South Asian (28.4%), European (10.2%) and Ashkenazi Jewish (1.7%, AJ) with the remaining ancestries each <1% (**Figure 1a**, “Mid. East” row). The large proportion of individuals with majority-South Asian genetic ancestry may arise from a technical limitation to strongly distinguish between Middle Eastern and South Asian ancestry when admixed with European ancestry.

Patients self-reporting as Ashkenazi Jewish had majority genetic ancestry coming from the Ashkenazi Jewish ancestry component for 80.2% of patients, from the European component for 16.8%, and from the Middle Eastern component for 2.7% of patients, with <1% with other majority-GAs (**Figure 1a**, “AJ” row). Since the AJ genetic ancestry distribution among the AJ is wide-ranging with a mode near 3/4 AJ genetic ancestry, those with majority-European genetic ancestry may still have substantial levels of AJ ancestry (**SI Figure 1**).

Self-reporting Northern Europeans had largely expected genetic ancestry, with 96.9% of Northern Europeans having majority genetic ancestry coming from the European component and 2.2% having majority-AJ genetic ancestry with the other majority-GAs at <1% of Northern Europeans (**Figure 1a**, “N. Eur.” row).

Self-reporting Southern Europeans also had less expected genetic ancestry compared to Northern Europeans, with 84.0% of Southern Europeans having majority genetic ancestry coming from the European component, 13.5% from the Middle Eastern component, and 1.5% from the AJ component with the other majority-GAs at <1% of Southern Europeans (**Figure 1a**, “S. Eur.” row).

For patients self-reporting as African or African-American, 90.7% had majority genetic ancestry from the African ancestry component (AF) and 6.3% had majority genetic ancestry from the European component (**Figure 1a**, “Afr. Am.” row). As the African ancestry distribution among this self-reported ethnicity has a mode at around 80% with a secondary mode at 40%, the subpopulation with majority-European ancestry is still expected to have substantial African genetic ancestry (**SI Figure 1**). A subpopulation with admixture between Middle Eastern and African ancestry was also observed; 2.2% of patients self-reporting as African or African-American had majority genetic ancestry from the Middle Eastern genetic ancestry component.

Three subpopulations were observed among those self-reporting as Hispanic: 1) those with European-Native American admixture, 2) those with European-African American admixture, and 3) those with admixture from European, Native American (NA), and African sources. In this analysis, only two-way admixture was considered and those with strong evidence for more admixture sources were excluded (described in Methods). The levels of European and Native American ancestry in group (1) were wide ranging, reflected in the observations that 72.1% of self-reported Hispanics had majority-European genetic ancestry and 24.4% had majority-Native American genetic ancestry (**Figure 1a**, “Hispanic” row). The African ancestry in group (2) was also wide ranging with a mode closer to 1/4th; 1.8% of Hispanics had majority-ancestry from the African ancestry component, yet many more had substantial African ancestry (**SI Figure 1**).

## References

1. Pletcher BA, Bocian M, American College of Medical G. Preconception and prenatal testing of biologic fathers for carrier status. American College of Medical Genetics. Genetics in medicine : official journal of the American College of Medical Genetics. 2006;8(2):134–135.

2. Prior TW, Professional P, Guidelines C. Carrier screening for spinal muscular atrophy. Genetics in medicine : official journal of the American College of Medical Genetics. 2008;10(11):840–842.

3. Grody WW, Thompson BH, Gregg AR, et al. ACMG position statement on prenatal/preconception expanded carrier screening. Genetics in medicine : official journal of the American College of Medical Genetics. 2013;15(6):482–483.

4. Committee on G. Committee Opinion No. 691: Carrier Screening for Genetic Conditions. Obstetrics and gynecology. 2017;129(3):e41–e55.

5. Hogan GJ, Vysotskaia VS, Beauchamp KA, et al. Validation of an Expanded Carrier Screen that Optimizes Sensitivity via Full-Exon Sequencing and Panel-wide Copy Number Variant Identification. Clinical chemistry. 2018;64(7):1063–1073.

6. Kaseniit KE, Collins E, Lo C, et al. Inter-lab concordance of variant classifications establishes clinical validity of expanded carrier screening. Clinical genetics. 2019;96(3):236–245.

7. Johansen Taber KA, Beauchamp KA, Lazarin GA, Muzzey D, Arjunan A, Goldberg JD. Clinical utility of expanded carrier screening: results-guided actionability and outcomes. Genetics in medicine : official journal of the American College of Medical Genetics. 2019;21(5):1041–1048.

8. Beauchamp KA, Johansen Taber KA, Muzzey D. Clinical impact and cost-effectiveness of a 176-condition expanded carrier screen. Genetics in medicine : official journal of the American College of Medical Genetics. 2019;21(9):1948–1957.

9. Committee on Genetics ACoO, Gynecologists. ACOG Committee Opinion. Number 325, December 2005. Update on carrier screening for cystic fibrosis. Obstetrics and gynecology. 2005;106(6):1465–1468.

10. Grody WW, Cutting GR, Klinger KW, et al. Laboratory standards and guidelines for population-based cystic fibrosis carrier screening. Genetics in medicine : official journal of the American College of Medical Genetics. 2001;3(2):149–154.

11. Edwards JG, Feldman G, Goldberg J, et al. Expanded carrier screening in reproductive medicine-points to consider: a joint statement of the American College of Medical Genetics and Genomics, American College of Obstetricians and Gynecologists, National Society of Genetic Counselors, Perinatal Quality Foundation, and Society for Maternal-Fetal Medicine. Obstetrics and gynecology. 2015;125(3):653–662.

12. Mersha TB, Abebe T. Self-reported race/ethnicity in the age of genomic research: its potential impact on understanding health disparities. Human genomics. 2015;9:1.

13. Li JZ, Absher DM, Tang H, et al. Worldwide human relationships inferred from genome-wide patterns of variation. Science. 2008;319(5866):1100–1104.

14. Genomes Project C, Auton A, Brooks LD, et al. A global reference for human genetic variation. Nature. 2015;526(7571):68–74.

15. Pritchard JK, Stephens M, Donnelly P. Inference of population structure using multilocus genotype data. Genetics. 2000;155(2):945–959.

16. Liu Y, Nyunoya T, Leng S, Belinsky SA, Tesfaigzi Y, Bruse S. Softwares and methods for estimating genetic ancestry in human populations. Human genomics. 2013;7:1.

17. Condit C, Templeton A, Bates BR, Bevan JL, Harris TM. Attitudinal barriers to delivery of race-targeted pharmacogenomics among informed lay persons. Genetics in medicine : official journal of the American College of Medical Genetics. 2003;5(5):385–392.

18. Dyson SM, Culley L, Gill C, et al. Ethnicity questions and antenatal screening for sickle cell/thalassaemia [EQUANS] in England: a randomised controlled trial of two questionnaires. Ethn Health. 2006;11(2):169–189.

19. Shraga R, Yarnall S, Elango S, et al. Evaluating genetic ancestry and self-reported ethnicity in the context of carrier screening. BMC Genet. 2017;18(1):99.

20. Wang C, Zhan X, Liang L, Abecasis GR, Lin X. Improved ancestry estimation for both genotyping and sequencing data using projection procrustes analysis and genotype imputation. American journal of human genetics. 2015;96(6):926–937.

21. Bray SM, Mulle JG, Dodd AF, Pulver AE, Wooding S, Warren ST. Signatures of founder effects, admixture, and selection in the Ashkenazi Jewish population. Proceedings of the National Academy of Sciences of the United States of America. 2010;107(37):16222–16227.

22. Lew RM, Burnett L, Proos AL, Delatycki MB. Tay-Sachs disease: current perspectives from Australia. The application of clinical genetics. 2015;8:19–25.

23. Henneman L, Borry P, Chokoshvili D, et al. Responsible implementation of expanded carrier screening. European journal of human genetics : EJHG. 2017;25(11):1291.

24. van der Hout S, Holtkamp KC, Henneman L, de Wert G, Dondorp WJ. Advantages of expanded universal carrier screening: what is at stake?European journal of human genetics : EJHG. 2016;25(1):17–21.

25. Ben-Shachar R, Svenson A, Goldberg JD, Muzzey D. A data-driven evaluation of the size and content of expanded carrier screening panels. Genetics in medicine: official journal of the American College of Medical Genetics. 2019;21(9):1931–1939.

